# Emergence and Evolution of Labor Pain Concern During Pregnancy: A Longitudinal Prospective Cohort Study

**DOI:** 10.64898/2026.01.29.26345142

**Authors:** Won Lee, Alexander J. Butwick, Pamela Huang, Abigail Wong-Rolle, Gabriel Salazar, Miriam Kuppermann

## Abstract

**Background:** The evolving nature of patient concerns during pregnancy and delivery, including labor pain, plays a key role in guiding antenatal counseling about labor and delivery, but the timing of these concerns has not been well characterized. Understanding when labor pain becomes a prominent concern for pregnant patients can inform the timing of antenatal education about labor analgesia.

**Objective:** This study aimed to determine how labor pain ranks among pregnancy-related concerns identified by nulliparous pregnant patients and how the relative prominence of these concerns changes across gestation.

**Method:** We conducted a prospective, single-center longitudinal cohort study of 53 English-speaking, nulliparous patients. Participants ranked their top seven concerns from a list of 13 pregnancy-related concerns, including labor pain, at 20, 24-, 28-, 32-, and 36-weeks’ gestation. Rankings were scored from 1 (lowest-ranked concern) to 7 (highest-ranked concern), with the six unranked concerns assigned a score of zero. Changes in labor pain concern scores over time were assessed using linear mixed-effects models, adjusting for maternal characteristics and pregnancy- and fetal-related complications.

**Results:** The score for concern about labor pain management increased with advancing gestational age, with mean adjusted rank scores increasing from 1.4 at 20 weeks to 3.8 at 36 weeks (*P* < 0.001). No demographic or clinical covariates were significantly associated with labor pain score. Peak scores were most commonly reported at 28- and 36-weeks’ gestation.

**Discussion:** While labor pain became a greater salient concern along pregnancy, for some nulliparous patients, concern about labor pain arose as early as 20^th^ week gestation, with significant individual variation in the timing of these concerns.

## INTRODUCTION

Labor pain is a multidimensional experience involving physical, emotional, and psychological elements, and decisions about its management are shaped by individual values and prior experiences.^1^ Patients’ satisfaction with their labor pain management may depend not only on the method of analgesia but whether their experience matches their expectations and preserves a sense of control over their course of labor.^2^ These elements may be shaped by the quality and timing of prenatal counseling.^3^ However, data are lacking on when pregnant patients develop interest about labor pain and management options in relation to other pregnancy- and delivery-related concerns.

Pregnant patients experience a range of pregnancy- and delivery-related concerns, including fetal health, maternal well-being, relationship dynamics, peripartum logistics, and financial pressures, in addition to labor experience and labor pain.^4,5^ The prioritization of these concerns may shift as pregnancy advances, often intensifying nearer to delivery.^6^ How labor pain ranks compared to other pregnancy- and delivery-related concerns is poorly described. Patients may first learn about labor analgesia only after arriving on the labor and delivery suite.^7,8^ This may lead to suboptimal decision-making about pain management due to the late timing of receipt of information, especially if patients are experiencing labor pain.

Understanding how and when labor pain concern emerges and evolves during pregnancy is critical for optimizing antenatal education and support, particularly for nulliparous parturients.^9^ Tailoring the timing of prenatal counseling to match patients’ evolving priorities can help close the gap between expectations and actual experience and support better-informed labor and delivery experiences.^10^ We, therefore, conducted a longitudinal study to examine how nulliparous pregnant patients’ ranking of labor pain concern changes over time during pregnancy compared to other pregnancy-related concerns.

## METHODS

This study was conducted in accordance with the ethical guidelines of the Declaration of Helsinki. The study was approved by the local Institutional Review Board (#22-37604) on 12/01/2022. All participants provided written informed consent prior to study enrollment. Study data are available upon request.

### Study Design

To identify pregnancy- and delivery-related concerns experienced during the antenatal period, we first conducted an exploratory preliminary study using semi-structured interviews with 15 nulliparous pregnant patients, purposively sampled to represent each trimester of pregnancy. Interviews were conducted by study investigators (WL and PH) via Zoom (San Jose, California), audio-recorded, and transcribed verbatim. An inductive thematic analysis was performed, with two independent coders (PH and AW) reviewing transcripts to identify emergent themes and discrepancies were adjudicated by a third reviewer (WL).^11^ Coding was conducted using Dedoose qualitative analysis software (Los Angeles, California). This process identified 13 distinct pregnancy-related concerns, including fetal well-being, postpartum planning, and labor pain (Table 1), which informed the development of the survey instrument.

**Table 1:**
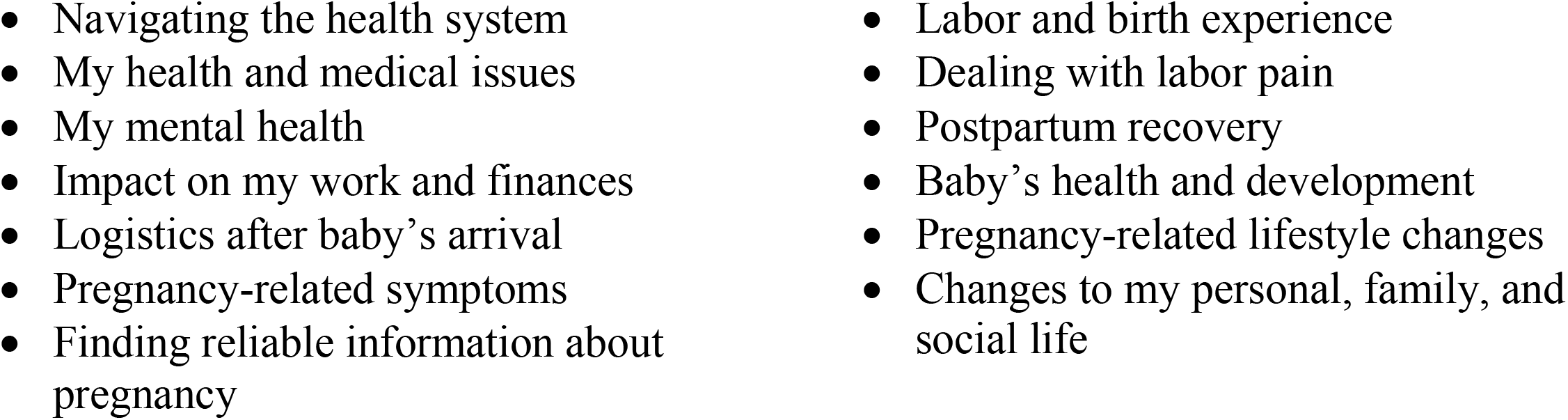
Thirteen pregnancy-related concerns identified during preliminary qualitative phase.

|  |  |
| --- | --- |
| • Navigating the health system<br>• My health and medical issues<br>• My mental health<br>• Impact on my work and finances<br>• Logistics after baby's arrival<br>• Pregnancy-related symptoms<br>• Finding reliable information about pregnancy | • Labor and birth experience<br>• Dealing with labor pain<br>• Postpartum recovery<br>• Baby's health and development<br>• Pregnancy-related lifestyle changes<br>• Changes to my personal, family, and social life |

In the main prospective longitudinal study, the 13 pregnancy-related concerns identified in the preliminary study were incorporated into a repeated-measures survey administered at five gestational time points (20-, 24-, 28-, 32-, and 36-weeks’ gestation) to evaluate how patients ranked these concerns and how their relative prominence compared to other concerns changed across pregnancy. Pregnant nulliparous patients with a gestational age of less than 20 weeks at enrollment who received prenatal care at the University of California San Francisco, a tertiary academic center, were eligible to participate. Patients were recruited between July 2023 and April 2024. Exclusion criteria included known major fetal anomalies at enrollment, planned transfer of care outside the institution, and lack of English language proficiency.

At each study time point, participants selected and ranked the seven concerns most important to them from a list of 13 concerns. Rankings ranged from 1 (most important) to 7 (least important), with the order of concern presentation randomized at each survey administration to minimize order effects. Ranked concerns were assigned scores from 7 (highest-ranked concern) to 1 (lowest-ranked concern), and unranked concerns received a score of 0, generating longitudinal rank-score data across five gestational assessments.

The survey was administered electronically via Qualtrics (Seattle, Washington) and designed for completion within 10–15 minutes. Patients were assigned unique study IDs to link survey responses across time points. They were asked to complete each survey within one week of the scheduled gestational time point, with two automated email reminders sent two days apart. Surveys not completed within the window were considered missing at that time point but did not disqualify patients from subsequent surveys. Study participants were remunerated up to $25 for completing all five surveys, with proportional compensation based on the number of surveys completed.

### Statistical Analysis

Descriptive statistics were used to summarize patient demographics and rank scores for pregnancy-related concerns at each gestational time point, with median and interquartile ranges (IQRs) reported. To analyze longitudinal changes in labor pain concern ranking and account for repeated measures, we conducted linear mixed-effects regression modeling, treating the rank score (0 - 7) as a continuous outcome. Maternal age, race (classified as White, Black, Asian, Latino/Hispanic, and Other), and new pregnancy or fetal complications, including gestational diabetes, hypertensive disorders of pregnancy, gestational thrombocytopenia, and intrauterine growth restrictions, that developed during the study period were included as fixed covariates. A random intercept for each patient accounted for individual baseline differences and for correlation among repeated measures. Model diagnostics included estimation of random effect variance and evaluation of model convergence. Beta coefficients with 95% confidence intervals (Cis) are reported. Margin analysis was performed to estimate model-adjusted mean scores and corresponding 95% CIs for each gestational week. Statistical significance was defined as two-sided *P* < 0.5. All analyses were conducted using Stata 16 (StataCorp, College Station, Texas).

In secondary analyses, we identified the time point at which each patient assigned their highest ranking to each concern (“peak score”). For tied peak scores, the earliest time point was used to reflect the first emergence of concerns. We applied a chi-square goodness-of-fit test to determine if the frequency of peak scores at each gestational week significantly deviated from a pattern expected under a uniform distribution. These analyses were limited to patients who completed all five survey time points.

## RESULTS

Fifty-three nulliparous pregnant patients enrolled in the prospective longitudinal study. Their demographic and clinical characteristics are summarized in Table 2. Nearly all participants (n = 50, 94%) completed all five surveys. Attrition was minimal and resulted from withdrawal at 28 weeks’ gestation (n = 1) or preterm delivery prior to the final assessment at 36 weeks (n = 2). Participants enrolled at a mean gestational age of 15.0 weeks and delivered at a mean gestational age of 38.8 weeks. Obstetric complications arising after enrollment were documented in 19 participants (36%), most commonly gestational diabetes (n = 8) and hypertensive disorders of pregnancy (n = 8). New fetal concerns developed in 11 participants.

**Table 2:** Participant Demographics and Clinical Characteristics.

| Characteristics | Overall (n = 53) |
| --- | --- |
| Mean Age Years (Standard Deviation) | 33.8 (3.7) |
| Race/Ethnicity (n, (%)) |  |
| White | 31 (58.5%) |
| Black | 1 (1.9%) |
| Asian | 16 (30.2%) |
| Latino/Hispanic | 1 (1.9%) |
| Other | 2 (3.8%) |
| Declined to answer | 2 (3.8%) |
| Insurance |  |
| Private | 53 (100%) |
| Gestational age at enrollment (weeks) (n, (SD)) | 15.0 (3.3) |
| Gestational age at delivery (weeks) (n, (SD)) | 38.8 (2.2) |
| Pregnancy-related complications (n, (%)) |  |
| Gestational diabetes | 8 (15.1%) |
| Hypertensive disorders of pregnancy | 8 (15.1%) |
| Gestational thrombocytopenia | 1 (1.9%) |
| Anemia | 7 (13.2%) |
| GBS<sup>+</sup> bacteriuria | 1 (1.9%) |
| Short cervix | 1 (1.9%) |
| Intrauterine growth restrictions | 3 (5.7%) |
| Macrosomia | 2 (3.8%) |
| Fetal deceleration in testing | 2 (3.8%) |
| Others | 4 (7.6%) |

<sup>+</sup>GBS = group B streptococcus

The median score for labor pain concern ranking rose progressively with gestational age, from 0 (IQR: 0–3) at 20 weeks to 5 (IQR: 1–6) at 36 weeks. In the linear mixed-effects regression model, compared to 20 weeks’ gestation, the labor pain concern rank score increased by 1.23 (95% CI: 0.43–2.02) points at week 28, 1.75 (95% CI: 0.95–2.54) points at week 32, and 2.36 (95% CI: 1.56–3.17) points at week 36 (Table 3). In the multivariable mixed-effects model, adjustment for maternal age, race, gestational age at enrollment, and new pregnancy or fetal complications produced no meaningful change in the week-specific coefficients, with adjusted estimates differing from the unadjusted values by less than 1% at each measured gestational age. The adjusted mean labor pain concern score increased from 1.42 (95% CI: 0.81 – 2.03) at 20 weeks, to 3.77 (95% CI: 3.15 – 4.40) at 36 weeks’ gestation (Figure A).

**Table 3:** Beta Coefficients and 95% Confidence Intervals for Labor Pain Scores during Pregnancy.

| **Gestational Age**<br>**(weeks)** | **Beta coefficient for**<br>**Labor Pain Score** | **95% Confidence**<br>**Interval** | **P value** |
| --- | --- | --- | --- |
| 20 | Reference |  |  |
| 24 | 0.51 | -0.28 – 1.30 | 0.207 |
| 30 | 1.23 | 0.43 – 2.02 | 0.003 |
| 34 | 1.75 | 0.95 – 2.54 | < 0.001 |
| 38 | 2.36 | 1.55 – 3.17 | < 0.001 |

**Figure A:**
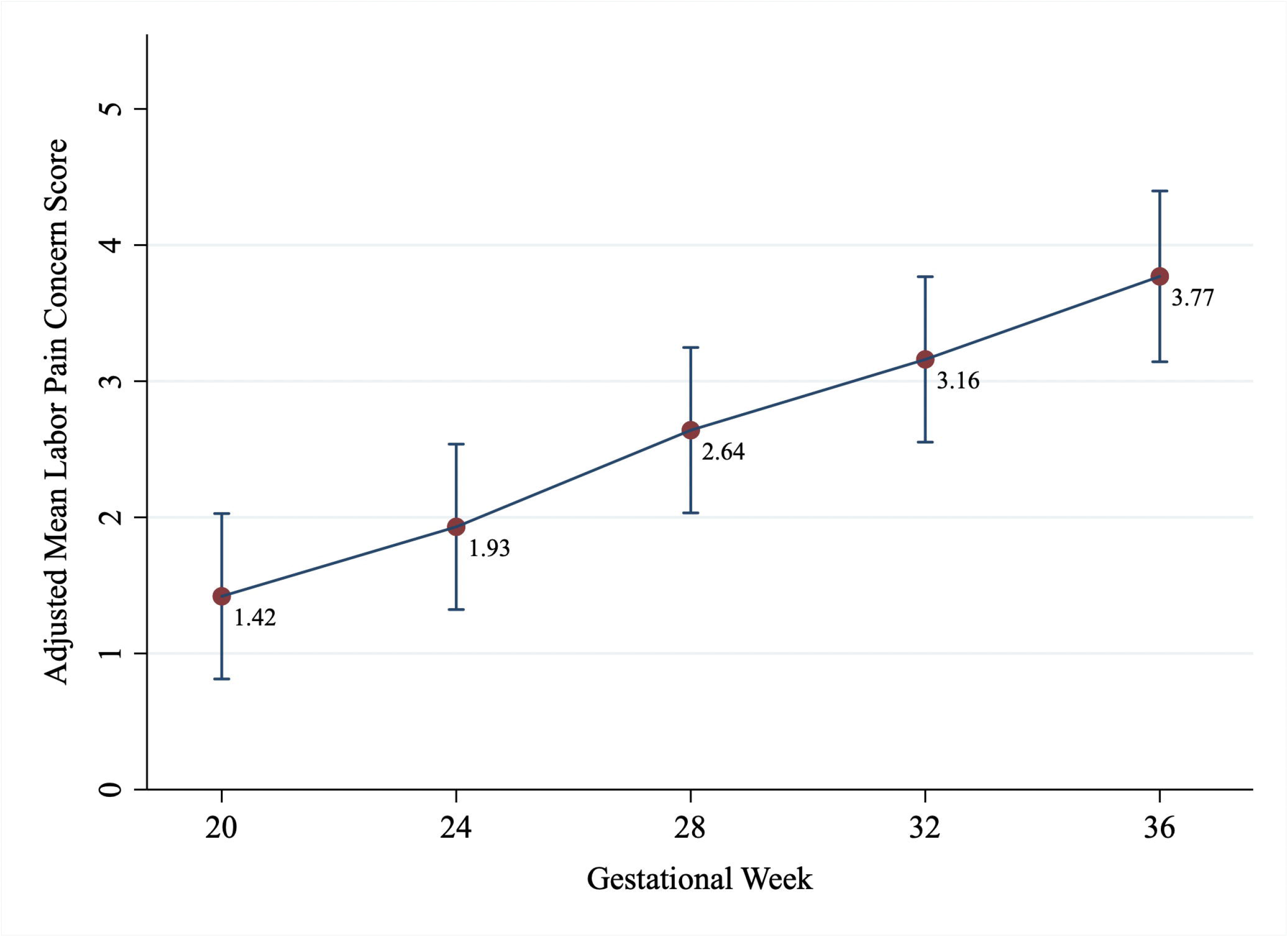
Adjusted Mean Labor Pain Concern Scores by Gestational Week. Adjusted mean labor pain concern scores by gestational week. Points represent model-based means of labor pain concern rank scores at 20, 24, 28, 32, and 36 weeks of gestation, estimated from a linear mixed-effects regression model, adjusted for age, race, gestational age at delivery, and new pregnancy and fetal complications. Error bars indicate 95% confidence intervals.

Among the 50 participants who completed all five surveys, the timing of the peak reported scores for labor pain are presented in Figure B. Peak scores were most commonly reported at 28 and 36 weeks. The frequency of peak scores for labor pain concern did not differ across gestational time points (*P* = 0.63).

**Figure B:**
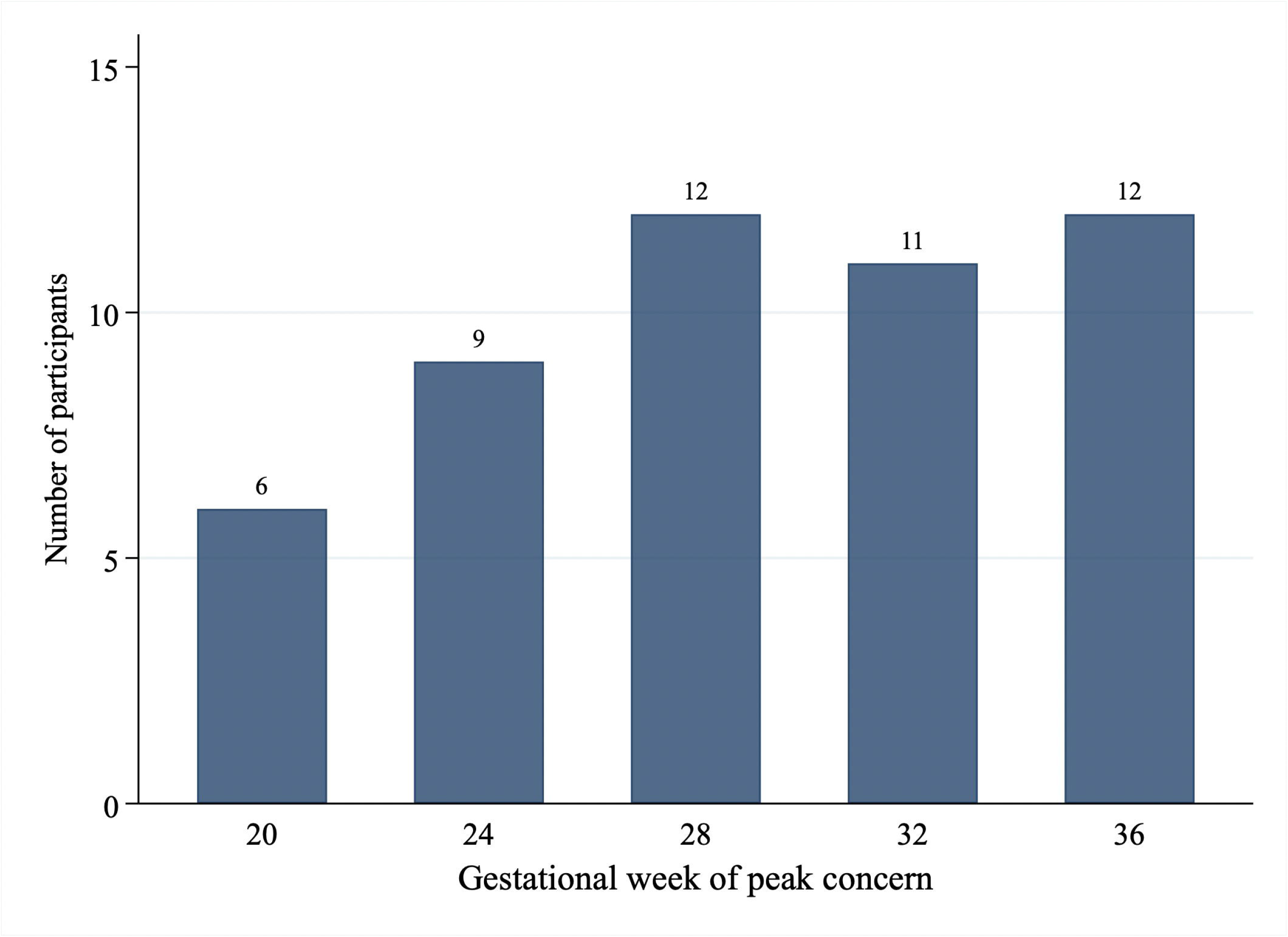
Distribution of Gestational Timepoints at Peak Labor Pain Concern. Bar plot showing the gestational week at which participants (n = 50) reported their highest level of concern about labor pain management. If multiple timepoints shared the same peak score, the earliest timepoint was used.

To contextualize labor pain concern among broader pregnancy-related concerns, we explored in secondary analyses how scores of all pregnancy, labor, and delivery concerns changed over time (Figure C). Compared to other pregnancy-related concerns, labor pain demonstrated an upward trajectory in prioritization. It was ranked 10th at 20 weeks, rose to 8th at 24 weeks, 4th at 28 weeks, 5th at 32 weeks, and reached 2nd at 36 weeks. The baby’s health and development was the top-ranked concern at 20 and 24 weeks but declined in priority by 36 weeks. Logistics after birth was the second-highest ranked concern at 20 weeks, rose to the highest concern at 28 and 32 weeks and was the third highest concern at 36 weeks. Labor and birth experience, a concern conceptually distinct from labor pain concern, rose from seventh place at 20 weeks to become the highest-ranked concern by 36 weeks.

**Figure C:**
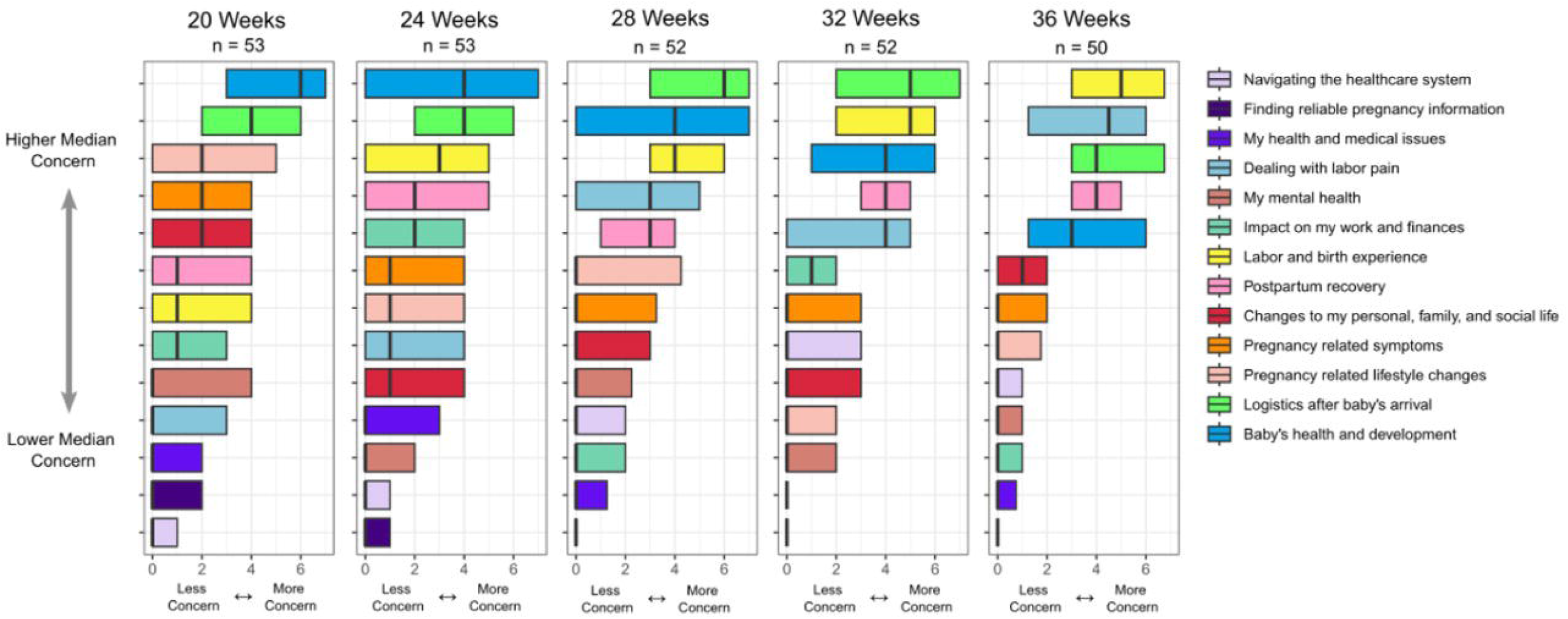
Ranking Trends of Key Pregnancy-related Concerns by Gestational Week. Boxplots of participant ranking of all 13 pregnancy-related concerns at each gestational timepoint. For each timepoint, concerns are ordered vertically by median rank score. Horizontally the box edges are the 25th and 75th percentile, with left being less concern and right being more concern.

## DISCUSSION

In this longitudinal study of nulliparous pregnant patients, we found that the mean rank score for labor pain concern more than doubled between 20 and 36 weeks, with the most prominent change observed between 20 and 28 weeks. Demographic and clinical factors, including age, race, and new pregnancy or fetal complications, did not explain the progressive increase in labor pain concern scores. Compared to other pregnancy-related concerns, labor pain ranking increased significantly from 10^th^ at 20 weeks’ gestation to second at 36 weeks’ gestation. These findings suggest that patients’ concern about labor pain increase substantially after 20 weeks and become a high priority in the third trimester.

The escalation in labor pain concern across gestational period that we observed aligns with previous studies showing that anxiety and fear about childbirth often intensifies as delivery approaches, especially for nulliparous patients.^12,13^ That the most pronounced rise in this concern occurred between 20 and 28 weeks may reflect a period when many patients transition from early prenatal concerns including fetal viability^14^ and significant physiologic stressors like hyperemesis^15^ to more concrete considerations about their upcoming labor and delivery. Given the high priority of labor pain concerns after 28 weeks’ gestation, our findings suggest that there is a potential need to address concerns about labor pain in the third trimester before admission to the labor and delivery unit.

Despite the overall upward trajectory in labor pain concern as pregnancy advanced, the timing of patients’ peak concern varied substantially. While most patients expressed elevated concern about labor pain after 28 weeks’ gestation, some reported high level of concern early in the second trimester. This variability may suggest that a one-size-fit-all approach to prenatal counseling may not fully address patient needs.^16^ By introducing discussions about pain management when patients are ready to engage or seek guidance, clinicians can give patients opportunities to consider their options and clarify preferences, which may enhance their birth experiences, expectations about labor pain, and decision-making about pain management.

Moreover, intervention studies are needed to assess whether tailoring the timing of counseling to meet evolving patient concerns can improve decision quality, patient satisfaction, or peripartum outcomes.^18^The early prominence of postpartum concerns further highlights the unpredictability of patient priorities. In our study, participants ranked postpartum logistics among their top concerns as early as 20 weeks’ gestation. Anticipatory guidance about the postpartum transition is often deferred until late pregnancy or even after delivery, yet our findings suggest that earlier engagement on these topics may be warranted. Pregnancy specific stress, such as concerns about parenting, postpartum transition, and additional financial burden of caring for a newborn, has been linked to postpartum depression, reduced early parent-infant bonding, and preterm birth.^19– 21^ Identifying when patients begin to experience these concerns may offer an important opportunity for timely discussion. Proactively addressing potential postpartum stressors early may support patient well-being and may mitigate downstream risks for both patients and infants.^22^

This study has several strengths. The survey response rate was high across all measured time points. The ranked concerns were developed with direct patient input from pilot interviews, enhancing the content validity of our measures. However, our study has several limitations. We conducted the study at a single academic center with English-proficient, nulliparous patients, limiting the generalizability of our findings. Additionally, while the ranking data allowed us to identify when labor pain became a more prominent concern, this approach does not capture the absolute intensity of concerns or subjective meaning behind patients’ ranking. Finally, we did not assess anxiety levels, mental health conditions, and other psychological indicators of well-being and how these influence pregnancy, labor, and delivery concerns.

In conclusion, we observed that, in a cohort of 53 nulliparous pregnant patients, labor pain concern increased after 20 weeks’ gestation and peaked in the third trimester. Further, in relation to other pregnancy concerns, the ranking of labor pain increased with advancing gestation and was the second-highest pregnancy-related concern at 36 weeks’ gestation. These findings provide early evidence about how patients’ concern and ranking of labor pain increases after 20 weeks. Evaluating whether counseling about labor pain and pain management in the third trimester influences patients’ decision-making about labor analgesia and their delivery experiences requires further investigation.

## Data Availability

All data produced in the present study are available upon reasonable request to the authors

## Acknowledgement

The authors thank Dr. Kiana Nguyen for assistance with the pilot phase of the study. We also thank Dr. Ronald B. George for early input on study design and conception.

## Reference

1 Labor S, Maguire S. The Pain of Labour. Rev Pain 2008;2(2):15. Doi: 10.1177/204946370800200205.

2 Anim-Somuah M, Smyth RMD, Cyna AM, Cuthbert A. Epidural versus non-epidural or no analgesia for pain management in labour. Cochrane Database of Systematic Reviews 2018;2018(5). Doi: 10.1002/14651858.CD000331.pub4.

3 Avalos LA, Oberman N, Gomez L, et al. Group Multimodal Prenatal Care and Postpartum Outcomes. JAMA Netw Open 2024;7(5):e2412280–e2412280. Doi: 10.1001/JAMANETWORKOPEN.2024.12280.

4 Hildingsson I, Larsson B. Women’s worries during pregnancy; a cross-sectional survey using the Cambridge Worry Scale in a rural area with long distance to hospital. Sexual & Reproductive Healthcare 2021;28:100610. Doi: 10.1016/J.SRHC.2021.100610.

5 Wilska A, Rantanen A, Botha E, Joronen K. Parenting Fears and Concerns during Pregnancy: A Qualitative Survey. Nurs Rep 2021;11(4):891. Doi: 10.3390/NURSREP11040082.

6 Peã’Acoba-Puente C, Monge FJC, Morales DM. Pregnancy worries: a longitudinal study of Spanish women. Acta Obstet Gynecol Scand 2011;90(9):1030–5. Doi: 10.1111/J.1600-0412.2011.01208.X.

7 Attanasio L, Kozhimannil KB, Jou J, McPherson ME, Camann W. Women’s experiences with neuraxial labor analgesia in the Listening to mothers II survey: A content analysis of open-ended responses. Anesth Analg 2015;121(4):974–80. Doi: 10.1213/ANE.0000000000000546,.

8 Wada K, Charland LC, Bellingham G. Can women in labor give informed consent to epidural analgesia? Bioethics 2019;33(4):475–86. Doi: 10.1111/bioe.12517.

9 Van Leugenhaege L, Degraeve J, Jacquemyn Y, Mestdagh E, Kuipers YJ. Factors associated with the intention of pregnant women to give birth with epidural analgesia: a cross-sectional study. BMC Pregnancy Childbirth 2023;23(1):598. Doi: 10.1186/s12884-023-05887-w.

10 Lally JE, Murtagh MJ, Macphail S, Thomson R. More in hope than expectation: a systematic review of women’s expectations and experience of pain relief in labour. BMC Med 2008;6(7). Doi: 10.1186/1741-7015-6-7.

11 Proudfoot K. Inductive/Deductive Hybrid Thematic Analysis in Mixed Methods Research. J Mix Methods Res 2022;17(3):308–26. Doi: 10.1177/15586898221126816.

12 Hildingsson I, Nilsson J, Merio E, Larsson B. Anxiety and depressive symptoms in women with fear of birth: A longitudinal cohort study. Eur J Midwifery 2021;5(August):1–9. Doi: 10.18332/EJM/138941.

13 Redondo MM, Liebana-Presa C, Pérez-Rivera J, et al. Exploring Self-Perceived Stress and Anxiety Throughout Pregnancy: A Longitudinal Study. Diseases 2025;13(4):121. Doi: 10.3390/DISEASES13040121.

14 Tong S, Kaur A, Walker SP, et al. Miscarriage risk for asymptomatic women after a normal first-trimester prenatal visit. Obstetrics and Gynecology 2008;111(3):710–4. Doi: 10.1097/AOG.0B013E318163747C.

15 Jarvis S, Nelson-Piercy C. Management of nausea and vomiting in pregnancy. BMJ 2011;342(7812). Doi: 10.1136/BMJ.D3606.

16 Raynes-Greenow CH, Roberts CL, McCaffery K, Clarke J. Knowledge and decision-making for labour analgesia of Australian primiparous women. Midwifery 2007;23(2):139–45. Doi: 10.1016/J.MIDW.2006.06.004.

17 Simona F, Sara B, Marta B, et al. Coping strategies for labor pain, related outcomes and influencing factors: A systematic review. Eur J Midwifery 2022;6(November):1–13. Doi: 10.18332/EJM/156440.

18 White E, Davies A, Demetri A, et al. Women’s perspectives of decision-making for labour and birth: a qualitative antenatal-postnatal paired interview study. BMJ Open 2025;15(6). Doi: 10.1136/BMJOPEN-2024-096171.

19 Armans M, Addante S, Ciciolla L, Anderson M, Shreffler KM. Resilience During Pregnancy: How Early Life Experiences are Associated with Pregnancy-Specific Stress. Advers Resil Sci 2020;1(4):295–305. Doi: 10.1007/S42844-020-00017-3/FIGURES/3.

20 Esparza KA, Shreffler KM. Stress across the perinatal period among low-income mothers and the moderating role of perceived social support. J Reprod Infant Psychol 2025. Doi: 10.1080/02646838.2025.2532594.

21 Katon W, Russo J, Gavin A. Predictors of postpartum depression. J Womens Health (Larchmt) 2014;23(9):753–9. Doi: 10.1089/JWH.2014.4824.

22 Matthey S. Anxiety and Stress During Pregnancy and the Postpartum Period. in: Wenzel A, editor. The Oxford Handbook of Perinatal Psychology, vol. 1. Oxford University Press; 2014. pp. 132–49.

